# Impact of small-area lockdowns for the control of the COVID-19 pandemic

**DOI:** 10.1101/2020.05.05.20092106

**Authors:** Cristóbal Cuadrado, María José Monsalves, Jean Gajardo, María Paz Bertoglia, Manuel Nájera, Tania Alfaro, Mauricio Canals, Jay S. Kaufman, Sebastián Peña

**Author notes:** **Corresponding Author:** Cristóbal Cuadrado, Escuela de Salud Pública, Universidad de Chile, Independencia 939, Independencia. **Funding**: The authors received no specific funding for this work.

## Abstract

**Background:** Countries confronting the COVID-19 pandemic are implementing different social distancing strategies. We evaluated the impact of small-area lockdowns in Chile, aimed to reduce viral transmission while minimizing the population disrupted. The effectiveness of this intervention on the outbreak control is unknown.

**Methods:** A natural experiment assessing the impact of small-area lockdowns between February 15th and April 25th, 2020. We used mobility data and official governmental reports to compare regions with small-area lockdowns versus regions without. The primary outcome was the mean difference in the effective reproductive number (Re) of COVID-19. Secondary outcomes were changes in mobility indicators. We used quasi-experimental methods for the analysis and examined the impact of other concurrent public health interventions to disentangle their effects.

**Results:** Small-area lockdown produced a sizable reduction in human mobility, equivalent to an 11.4% reduction (95%CI −14.4% to −8.38%) in public transport and similar effects in other mobility indicators. Ten days after implementation, the small-area lockdown produced a reduction of the effective reproductive number (Re) of 0.86 (95%CI −1.70 to −0.02). School and university closures, implemented earlier, led to a 40% reduction in urban mobility. Closure of educational institutions resulted in an even greater Re reduction compared with small-area lockdowns.

**Conclusions:** Small-area lockdowns produced a reduction in mobility and viral transmission, but the effects were smaller than the early closures of schools and universities. Small-area lockdowns may have a relevant supporting role in reducing SARS-CoV-2 transmission and could be useful for countries considering scaling-down stricter social distancing interventions.

## Introduction

The first case of COVID-19 in Chile was officially reported on March 3th(1), spreading rapidly along the country during the following weeks. (2) While on March 11th the World Health Organization (WHO) described the situation of COVID-19 as a global pandemic, in Chile several clusters of COVID-19 were declared on March 14th. (3,4) By then, many countries had implemented large-scale subnational lockdowns (e.g. Wuhan, Lombardy). (5,6) On March 15th, the Chilean government ordered the mandatory closure of schools, followed by the announcement of voluntary university closures and reorganization for remote instruction. (7)

On March 18th, the Chilean government declared a 90-day State of Emergency, enabling exceptional measures to limit certain rights or constitutional guarantees such as free transit. Starting March 19th, the government ordered the closure of all the country’s shopping centers. (8) The Chilean Medical Association and several other leading organizations urged the government for stronger measures and called for the population to stay at home (9). With this and the active coverage of mass media, the urgency of the COVID-19 pandemic became more evident to the population, increasing broader awareness. In the following weeks, the Government implemented several additional public health interventions, including a ban on mass gatherings and an overnight curfew starting on March 22nd, among others. Furthermore, this prompted the application of a teleworking regime in most offices with the capacity for remote operations.

One of the main responses from the national health authorities throughout April 2020 was the policy of small-area lockdowns. These are general quarantines applied to small areas, usually municipalities or even neighborhoods, forcing the population within them to stay at home with the exception of medical emergencies or purchases of food and essential medicines. Continuity of this measure is evaluated every 7 days, according to the cumulative incidence of cases and other indicators. (10) On March 26th, the first lockdown was declared on seven municipalities of the Metropolitan Area. To our knowledge, Chile is the only country worldwide officially implementing such small-area lockdowns.

Small-area lockdowns aim to reduce viral transmission while minimizing the size of the population that needs to be disrupted to achieve this end. This approach could be more easily enforced and feasible to be implement in certain context such as Chile, considering an ongoing period of national civil unrest (11), were a large-scale national lockdown could have been unpalatable to the populace difficult to sustain politically. Additionally, they provide an alternative solution to transition in a controlled way to normality as the outbreak evolves. Despite the emerging evidence of general lockdowns, (12) it is unclear whether these interventions focusing on smaller areas are effective in modifying population behaviors, such as spatial mobility, and reducing the incidence of new cases.

To explore this, we exploit the implementation of small-area lockdowns during the early development of the COVID-19 pandemic in Chile to assess their impact on mobility and case transmission.

## Methods

### Study design and setting

We evaluated the effects of a natural experiment(13) on changes to human mobility and case transmission due to small-area lockdowns in response to the COVID-19 pandemic, from February 15 to April 15, 2020 in Chile. We employed quasi-experimental methods for this aim. We report the study in accordance with the Strengthening the Reporting of Observational Studies in Epidemiology (STROBE) guidelines(14,15).

### Variables

#### Public health interventions

Our main intervention of interest is the implementation of small-area lockdowns in Chile. The lockdowns started in the Metropolitan Region and expanded to 6 regions by April 15 th. People in locked-down areas are mandated to stay at home, allowed to leave for a period of two hours twice a week for essential purposes such as seeking medical care or purchasing foods and basic goods (16). Since other public health interventions are also implemented during the study period, we included them in our analysis to disentangle their distinct effects. These concurrent interventions include the closures of schools and universities that started nationally on March 15^th^, and the overnight curfew (from 12 am to 5 am) implemented nationally on March 22 continuing up to the present time.

#### Comparator

Intervention and analysis are at the level of the region (i.e. 16 in Chile). Regions are the first-level administrative division in Chile. Comparators are regions without small-area lockdowns in the study period, but where schools were closed, and an overnight curfew was implemented. Details on assignment of regions to intervention or control conditions are shown in the Supplemental Appendix (S1).

### Outcomes

#### Reproductive number

The primary outcome is the effective reproductive number (Re) as an indicator of case transmission over the epidemic. We estimate the Re using the method developed by Cori et al (2020)(17) considering the case incidence during the last week (7 days) for each region and a serial interval *τ* = 5 days(18) with usual variability between 3 and 7 days.(19,20)

#### Human mobility

The secondary outcome is human mobility, which is a surrogate for the effect of public health interventions and a potential mediator between the small-area lockdown intervention and the expected effect on the Re. We include four indicators of human mobility: (1) public transport, hubs and subway, bus, and train stations; (2) Mobility for retail and recreation such as to restaurants, cafes, shopping centers; (3) Mobility to the workplace and; (4) Residential mobility.

We report proportional changes in human mobility between February 15th and April 25^th^, 2020. The changes for each day are compared with the median value for the corresponding day of the week, during the 5-week period Jan 3–Feb 6, 2020.

### Data sources

We used data from three information sources: Google Mobility Data, for mobility indicators;(21) official reports by the Ministry of Health of Chile on daily incident cases of COVID-19(4); and, official decrees by the Chilean government for lockdown dates.

### Statistical Analyses

#### Effects over human mobility

We conducted a difference-in-difference analysis to estimate the effects of the small-area lockdown in the intervention regions versus control regions. For this purpose, we used a linear regression model specified as follows:

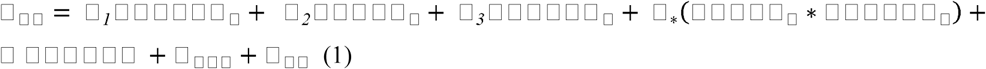

where 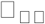 is the mobility outcome for the region i at the day t. “School” is the indicator variable for school and university closures at time t (identical for all sites), “Group” is the indicator variable for the regions where a lockdown is implemented, taking the value 1 for lockdown sites and 0 for controls. “Period” is a categorical variable with five levels that represent the periods of lockdown implementation: 1) no lockdown in effect; 2) partial lockdown of the Metropolitan Region; 3) lockdown of Temuco city (Araucanía Region); 4) lockdown of the cities of Chillán (Ñuble Region) and Osorno (Los Lagos Region); 5) lockdown of Punta Arenas City (Magallanes Region). This variable captures the time fixed-effects of each period. To control for seasonality, we used a vector of day dummy variables represented by *δ_day_*. Additionally, we adjusted for a time-fixed effect (*η*) to capture the effect of Easter holiday. The idiosyncratic error is represented by *ε_it_*. Since we have five implementation periods, the interaction with the group variable produces a vector of four coefficients of interest (_*_) for difference-in-difference estimation. These coefficients capture the impact of lockdowns on human mobility at different levels of implementation compared with the no lockdown period and among regions that never experienced lock-down.

#### Effects over case transmission

We estimate a similar difference-in-difference model to assess the impact of public health interventions on the effective reproductive number (Re) using a model similar to Equation 2 but considering a 10-day lag between intervention and the outcome variable. This lag was chosen to account for the expected time between contagion and case confirmation: 5 days for pre-symptomatic phase(22) and 5 days between first symptom and confirmation based on official reports of the Chilean Ministry of Health(23). Additionally, a model to assess the mediator effect of human mobility on the Re was estimated. Details on this model are available in the Supplemental Appendix (S2), Results for the effect over Re are reported in absolute values, but also as a relative reduction using as reference the R0 estimated for the SARS-CoV-2(24).

To account for the uncertainty of the time between exposure and outcome, we explore different lags between 7 and 14 days in a sensitivity analysis. Results are available in the Supplemental Appendix (S3). We also explore the robustness of our results using alternative models based on instrumental variables (S3)

For all models robust standard errors are used to compute 95% confidence intervals. All analyses were implemented in R version 3.6.2 (R Foundation for Statistical Computing). Dataset and R script are available in the Supplemental Appendix (S4).

## Results

We observed changes in all mobility indicators, with a breakdown concomitant to school and university closures. The most evident decrease was observed in recreational mobility, followed by public transport and workplace mobility. After school and university closures, there was a sustained change in mobility, establishing a new pattern, with close to household movements. This pattern remained relatively stable during the following weeks, observing a second decrease over a statutory holiday (Easter weekend) at the end of the second week of April (Figure 1).

**Figure 1:**
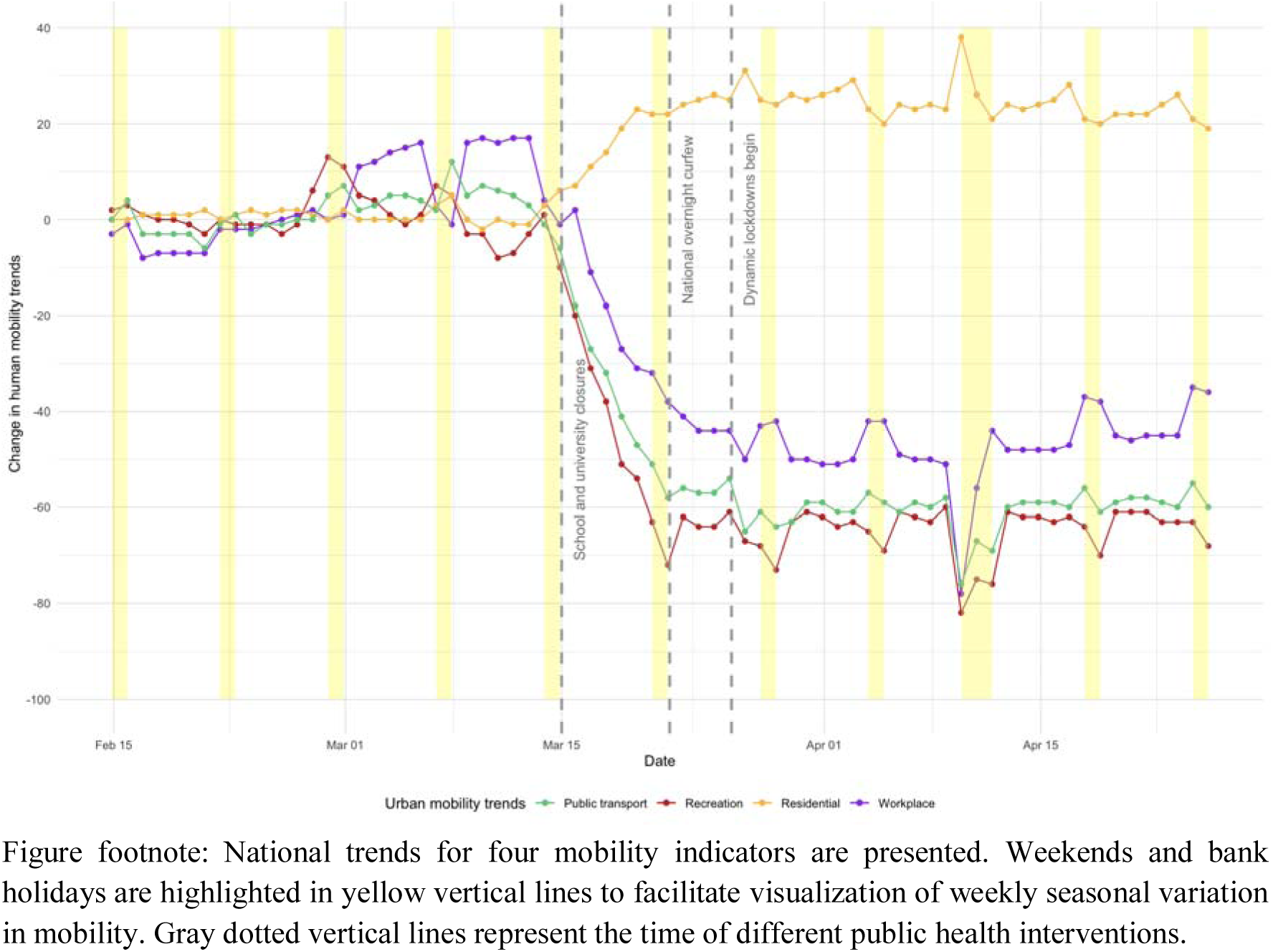
National trends in human mobility in the context of COVID-19 pandemic in Chile

Before the small-area lockdowns, intervention and control regions showed parallel trends (i.e. the parallel trend assumption in DiD was met) (Figure 2). Intervention regions showed lower workplace, retail, and public transport mobility compared with control regions. The difference between intervention and control regions widened following the implementation of the lockdown in the city of Temuco.

**Figure 2:**
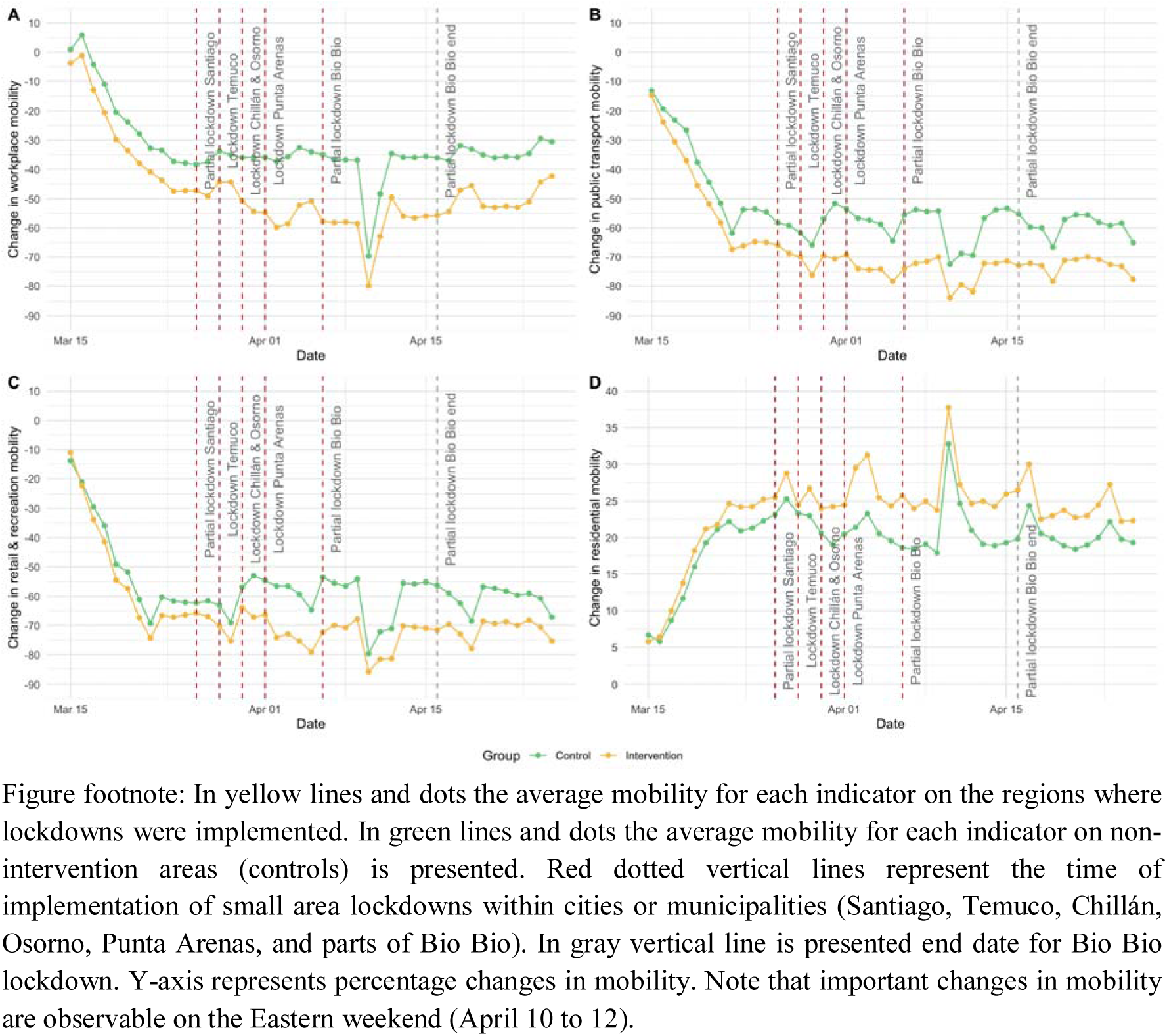
Trends in human mobility in regions with and without small-area lockdowns

Table 1 shows the result of the DiD analyses. We observed an important effect of the closure of schools and universities on mobility indicators, with the greatest effect observed for retail mobility (-45.7%, 95% CI −48.6 to −42.8) and workplace mobility (−40.5%, 95% CI −43.5 to −37.1). Small area lockdowns produce a further reduction by nearly 12% of the human mobility, particularly when the policy was already under effect in several areas. This effect occurs on top of the already important impact of school and university closures, substantially reducing human mobility.

**Table 1:**
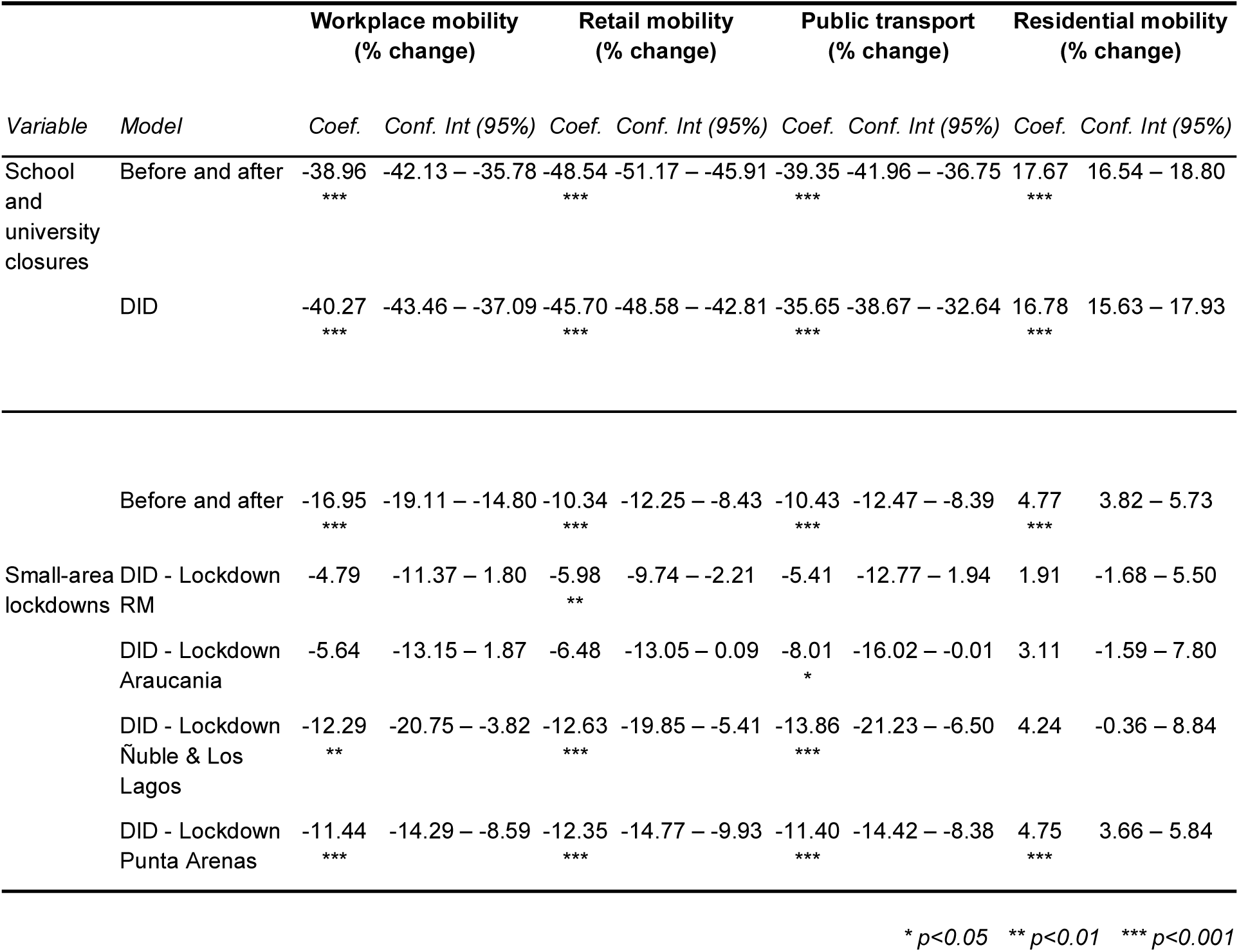
Impact of small-area lockdowns on human mobility

Results of models estimating the effect of small-area lockdowns over disease transmission are summarized in Table 2. All models suggest a reduction in the Re following small area lockdowns. Our estimates from the DiD model indicate a reduction of 0.86 (CI 95% 1.70 to −0.02) in the Re, evident only in the later phase of implementation when 9 municipalities (representing 10.2% of the national population) have a lockdown under effect. This effect is equivalent to a 34.4% reduction of the reported R0 for the disease. Additionally, school and university closures produced an even greater absolute reduction equivalent to a 2.03 (CI 95% −2.76 to −1.29) reduction in Re.

**Table 2:** Impact of small-area lockdowns on the effective reproductive number (Re)

| *Variable* | Before and after model |  | DID model |  |
| --- | --- | --- | --- | --- |
|  | *Estimates* | *Conf. Int (95%)* | *Estimates* | *Conf. Int (95%)* |
| School closure | -2.38 *** | -3.05 – -1.71 | -2.03 *** | -2.76 – -1.29 |
| Small-area lockdowns | -0.38 | -0.93 – 0.17 |  |  |
| Lockdown RM |  |  | -0.81 | -1.78 – 0.15 |
| Lockdown Araucania |  |  | -0.73 | -1.64 – 0.18 |
| Lockdown Ñuble & Los Lagos |  |  | -0.63 | -1.52 – 0.26 |
| Lockdown Punta Arenas |  |  | -0.86 * | -1.70 – -0.02 |
| Observations | 660 |  | 660 |  |
| R<sup>2</sup> / R<sup>2</sup> adjusted | 0.360 / 0.336 |  | 0.338 / 0.319 |  |

\* $p < 0.05$ \*\* $p < 0.01$ \*\*\* $p < 0.001$

Our estimates of the effect of human mobility over disease transmission suggest that for each 1% reduction in workplace or retail & entertainment mobility, the Re is expected to decrease by about 0.04 (S2 - Effects of human mobility of the effective reproductive number). Our best estimate of the impact of the small area lockdowns on human mobility is nearly an 11.4% reduction, therefore at most a 0.46 reduction of the Re mediated through human mobility could be expected, nearly 70% of the total effect detected.

Sensitivity analysis based on alternative quasi-experimental methods are presented in the Supplemental Information (S3 - Sensitivity analysis). We conducted IV models that confirm our DiD results, with slightly smaller effect sizes of the small-area lockdown over Re (−0.66; CI 95% −0.91 to −0.42). Additionally, we assessed the robustness of our results to different time-lags between intervention and outcome. Our conclusion remain unchanged to changes in this assumption between 7 and 14 days, with small area lockdowns producing a reduction of Re between −0.96 (CI 95% −1.89 to −0.03) and −0.83 (CI 95% −1.59 to −0.07) respectively for the DID model or between −0.88 (CI 95% −1.13 to −0.63) and −0.58 (CI 95%(−0.82 to −0.35) for the IV model.

## Discussion

This study showcases the important effects of public health interventions on human mobility and epidemic transmission. We study the effect of a seemingly unique policy implemented in Chile: small-area lockdowns. Our study suggests that small areas lockdowns could contribute for controlling the outbreak. Although an effect of a 0.4 reduction in the Re is small compared to the impact of other interventions, they may play a key-role as a coadjuvant for containment, once the decline has been established by other public health interventions, suiting it particularly well as an exit strategy for large-scale lockdowns.

Human mobility may play an important role in the early stage of the COVID-19 epidemic (25). Concordantly, our study shows an important reduction of mobility and concordant effects on the epidemic spread after the implementation of school and university closures. This effect should be understood beyond its obvious attributable impact on the reduction of transmission among school and young people, but as a modulator of urban mobility, adding more dynamic and complex pathways for the impact of social distance interventions. It seems likely that the early closure of schools and universities had a significant impact on the reduction of SARS-CoV-2 transmission during the first stage of the pandemic in Chile.

Different countries have implemented lockdowns, although their impact on the spread and on the economy is still to be seen. However, experiences on Spanish flu in 1918 showed different trends in the epidemic morbimortality according to the strictness of the social distancing measures implemented, with better results for cities with earlier and more restrictive interventions (26). While some countries are starting to bring their epidemic outbreaks under control, evidence on the effectiveness of current lockdowns is only now starting to come into focus (5,27–29). In Wuhan, China, the growth rate of the cases decreased after the implementation of a large-scale lockdown which included travel restrictions, quarantine and social distancing measures like closing of schools (27,30). In Huangshi the epidemic curve became flattened 9 days after a lockdown was implemented (27). Nevertheless, the most adequate strategy for implementing a lockdown (scale and timing) is still a matter of controversy (31). Interestingly, our study adds to the literature that small-areas lockdowns could also contribute as secondary measures for controlling the outbreak. Moreover, they could be particularly useful as a way out of large-scale lockdowns, after the epidemic outbreak was been already controlled.

Previous studies have estimated that school closure can reduce the size of an influenza outbreak by 13-25% (32,33). However, in the context of the COVID-19 pandemic, early evidence has been less auspicious given the lower transmission observed in children (34). Based on mobility data, our results show that the closure of schools and the limitation of the attendance of teachers, workers, and students to educational centers had an impact on the reduction of disease transmission. We hypothesize that this probably occurs because the closure is associated with other social processes that affect mobility within cities and caused the acceleration in the adoption of other measures such as the installation of work from home, as seen by other authors (35), and other individual actions of voluntary confinement. Individual voluntary quarantine decisions resulting from the increasing awareness are likely to have an effect on this outcome, but probably varying substantially depending on material circumstances and other socioeconomic considerations. However, due to the magnitude and evident temporal relationship with the closure of schools and universities, the effect of this measure seems evident.

Our study has a number of limitations. First, the lockdowns are implemented at a small-area level, but our analysis was aggregated at the regional level. While this can dilute the intervention effect, it seems adequate given that mobility in major cities occurs between different municipalities. Moreover, policy effects are expected to impact disease transmission on a larger scale and not only in the small area directly affected by the lockdown. Secondly, our main outcome (Re) is subject to biases due to underreporting or different testing capacity between regions, urban mobility indicators are not. Even so, the consistency of the results over mobility indicators, not affected by such bias, and the case transmission reinforce our conclusions. Third, our small-area lockdowns effect estimates could be underestimated due to the impact of previously implemented policies. Reducing mobility even more after archiving already low levels of movement is probably difficult. Fourth, our assessment of school and university closures is based on a before and after model, no control was available since all regions implemented the intervention on the same date. Therefore, it is not possible to disentangle the effects over mobility produced by school closures and other concomitant factors such as increasing awareness and observed effects cannot be entirely attributed to the closures measure. Fifth, Google mobility data used in this study can be biased towards groups with greater access to smartphones (36). In Chile 85% of the population has a mobile phone, with a slight gradient between socioeconomic groups (93% in high-income and 77% in low-income households) (37). Even when this data has been used to adequately infer human mobility in other contexts (38), our conclusions do not necessarily capture the changes of mobility patterns of more vulnerable groups.

Our study highlights the usefulness of mobility data for real-time surveillance of public health interventions, allowing national authorities to anticipate the effects over the epidemic curve. Therefore, it is advisable to include monitoring changes in mobility patterns as part of this point in the key information criteria of the outbreak progress. An important challenge for the Chilean context is to maintain the low trend of urban mobility observed until mid-April with its consequent effect on maintaining a controlled outbreak.

A small-area lockdown strategy requires good surveillance capacity, early warning alerts and the capacity to mobilize rapidly actions in case of an increase in the case transmission. If these conditions are not fulfilled, such a strategy is prone to fail. Since this is a short-term evaluation, it is still to be seen if such social distancing approach can be maintained and show sustained effectiveness in the mid-term to keep the epidemic curve under control. Based on the response seen in Chile, lifting school closures too early could produce an increase in urban mobility that could impact on disease transmission. Therefore, any action in this direction needs to be carefully planned to mitigate the risk of an outbreak rebound.

## Conclusion

Small-area lockdowns implemented in Chile have contributed to decreasing COVID-19 epidemic transmission, in the context of other major public health interventions. This quasi-experimental study contributes to reduce the evidence gap about the effectiveness of small-area lockdowns in the control of the COVID-19 pandemic. The achieved reductions in case transmission, although relatively smaller in magnitude compared with other interventions, could be useful for countries planning on lifting restrictions to maintain outbreak control while minimizing the social and economic impacts of larger-scale lockdowns.

## Data Availability

Data and code will be available in the supplemental appendix of the final manuscript

